# A Viral Fragmentation Signature for SARS-CoV-2 in Clinical Samples Correlating with Contagiousness

**DOI:** 10.1101/2021.01.11.21249265

**Authors:** Yukti Choudhury, Chae Yin Cher, Zi Yi Wan, Chao Xie, Jing Shan Lim, Ramandeep Kaur Virk, Min Han Tan, Alvin Kuo Jing Teo, Li Yang Hsu

## Abstract

The viral load of SARS-CoV-2 in clinical samples as measured by the primary diagnostic tool of RT-PCR is an imperfect readout for infection potential as most targeted assays designed for sensitivity, indiscriminately detect short and long RNA fragments, although infectivity is embodied only in the whole virus and its intact genome. Here, we used next-generation sequencing (NGS) to characterize 155 clinical samples and show sensitive and quantitative detection of viral RNA which confirmed subgenomic RNA in 57.6% of samples and provided a novel method to determine relative integrity of viral RNA in samples. The relative abundance of long fragments quantified as a viral fragmentation score was positively associated with viral load and inversely related to time from disease onset. An empirically determined score cut-off for presence of substantially fragmented RNA was able to identify 100% of samples collected after 8 days of illness with poor infection potential in line with current clinical understanding of infectiousness of SARS-CoV-2. The quantification of longer fragments in addition to existing short targets in an NGS or RT-PCR-based assay could provide a valuable readout of infection potential simultaneous to the detection of any fragments of SARS-CoV-2 RNA in test samples.

## Introduction

In March 2020, the World Health Organization (WHO) declared a pandemic of the coronavirus disease 2019 (COVID-19), an infection caused by severe acute respiratory syndrome coronavirus 2 (SARS-CoV-2)^1^. By end of 2020, 82 million infections have been reported worldwide with 1.79 million deaths (https://coronavirus.jhu.edu). Diagnostic tools for detection of present and past infection of SARS-CoV-2 have been crucial for the management of this public health emergency, and include an array of molecular tests and immunoassays^2^. Direct detection of nucleic acids via reverse-transcription-polymerase chain reaction (RT-PCR) is the most widely deployed modality. Next-generation sequencing (NGS) is a related molecular method, and is characterized by the ability to identify polymorphisms and to define interrelatedness of virus strains^3,4^, facilitated by the whole-genome interrogation of the virus.

Sensitive methods for detecting SARS-CoV-2 have been paramount in the identification and isolation of infected individuals and their close contacts, reducing viral transmission. Releasing infected persons prematurely from isolation carries the risk of fuelling transmission, while prolonging isolation unnecessarily can adversely affect resource allocation and cause undue disruptions^5^. While RT-PCR assays are widely regarded as the most sensitive of diagnostic methods, the inability to distinguish intact viral RNA – packaged in virions and able to cause onward transmission – from products of degradation of viral RNA ineffectual at further infection, has led to considerable debate on the dichotomous interpretation of RT-PCR results as positive/negative, and the in-depth analysis of the period of infectiousness^6,7^. Positive detection of virus RNA does not prove presence of infectious virus, as persistent detection of viral RNA in samples has been reported up to 35 days from symptom onset^8–12^, and after resolution of clinical symptoms of both mild and severe disease^13–16^. As large-scale vaccination is deployed across countries^17–19^, the uncertain impact of vaccination on potential contagiousness^20,21^, further highlights the need for means to quantify contagiousness accurately.

The isolation of whole virions in cultured cells, in contrast to RNA fragments provides evidence of the isolate’s replicative potential and more closely reflects the true infection potential^6^. Duration of live virus detection is much shorter than viral shedding and no live virus has been isolated in samples taken more than 8 days since symptom onset in immunocompetent patients ^8,11,22^. In accordance with this, in a hamster model, transmissibility of SARS-CoV-2 correlated with detection of infectious virus by culture, but not by RT-PCR-positivity alone, and the communicable period was less than 6 days for inoculated donor animals^23^. Viral load dynamics also differ between upper respiratory tract samples, where highest viral loads were reported at the time of symptom onset or soon after at 3-5 days of illness followed by consistent decline, and in lower respiratory tract samples where peak viral load is in the second week of illness^7^. It is at present unclear if this dynamic is related to dynamic infection potential of transmitted material from each site.

Detection of subgenomic RNA (sgRNA) has been used as a proof of viral replication, which can only happen in infected cells, and declines over days 10-11 from infection^8^. sgRNA does not get packaged into virions therefore detection of sgRNA is indicative of actively infected cells in a sample, and complements the detection of infectious viral RNA packaged into virions. SgRNA have been detected in cultured cells and in limited clinical samples most commonly by NGS and by RT-PCR for specific sgRNA fusion products, but are not routinely studied ^8,24,25^.

Prompted by the limitations of the most commonly used diagnostic device for SARS-CoV-2 for informing contagiousness, and by the urgent need to address this crucial parameter, in this study we employed whole-genome NGS with molecular barcoding and sequencing error-correction to characterize clinical samples collected from a cohort of migrant workers in Singapore. As a comprehensive tool with full genome targeting, NGS can inform presence of any SARS-CoV-2 RNA, the presence of sequence variants and sgRNA, and as we show for the first time, this method can also characterize the lengths of fragments of RNA in a sample. We go on to show that fragment lengths, characterized as a viral fragmentation score, are related to intactness of viral RNA, which is a prerequisite for infectiousness in the form of whole virus, and is correlated in with other clinical parameters of potential infectiousness such as viral load, sgRNA and duration of illness.

## Materials and Methods

### Sample collection

Samples used in this study were collected as part of a previous study conducted in a large cohort of migrant workers in Singapore, and evaluated and approved by the Director of Medical Services, Ministry of Health, under Singapore’s Infectious Disease Act^26^. A total of 155 samples, corresponding to 48 individuals were analyzed by NGS. The samples included 37 nasopharyngeal (NP) swabs collected in 3 mL of viral transport medium, 60 self-nasal swabs, and 58 oropharyngeal saliva samples collected in 2 mL of viral RNA stabilization fluid (SAFER™ VTM, Lucence, Singapore). The majority of the cases (35 of 48, 72.9%) were symptomatic at the time of first sample collection, one case was presymptomatic at time of first sampling, and 12 cases (25%) were asymptomatic through the study. For asymptomatic cases, the day of the first diagnostic test was used to determine days since diagnosis. For 17 cases (35.4%), only a single sample was available for sequencing, and for the remaining 31 (64.5%) of cases, between 2 and 9 samples were studied. For 5 cases (10.41%), samples from different sites (NP, nasal swab or saliva) were available but from the same timepoint (not longitudinal). All samples had previously been characterized for presence of SARS-CoV-2 using an RT-PCR assay using primers and probes sequences from the CDC 2019-nCoV Real-Time RT-PCR Diagnostic Panel^26,27^, henceforth referred to as CDC RT-PCR assay. Samples were selected for NGS based on SARS-CoV-2 positivity at at least one of 2 or 3 timepoints of sample collection and relatively high viral loads as estimated by the RT-PCR assay. Archived extracted RNA from anonymized samples were used for this study.

### Sample processing

Viral nucleic acid is extracted from the specimens (200 or 400 μl) using Viral Nucleic Acid Extraction Kit II (Geneaid Biotech Ltd., Taiwan) or QIAsymphony DSP Virus/Pathogen Kit (QIAGEN). For each sample, aliquots of extracted nucleic acid (RNA) was screened for presence of SARS-CoV-2 RNA using real-time reverse transcription-polymerase chain reaction (RT-PCR) targeting the N1 and N2 markers as specified by Centers for Disease Control and Prevention (CDC), and also processed for library preparation for NGS. Samples were processed in a College of American Pathologists (CAP) accredited and a Clinical Laboratory Improvement Amendments (CLIA) licensed laboratory (Lucence).

### Analytical validation of SARS-CoV-2 NGS method

Lower limit of detection (sensitivity) of the NGS method was determined using synthetic SARS-CoV-2 RNA spiked into pooled clinical matrix that tested negative for the virus by a RT-PCR method. For the analytical validation of NGS, synthetic SARS-CoV-2 RNA genomes (Twist Bioscience, San Francisco, USA) of known concentrations were spiked in to pooled negative clinical matrix (nasopharyngeal swab specimens) at known copy numbers prior to RNA extraction. The synthetic controls corresponded to these GenBank IDs of various published SARS-CoV-2 genome isolates. MT007544.1 (SKU: 102019), MN908947.3 (SKU: 102024), LC528232.1 (SKU: 102860), MT106054.1 (SKU: 102862), MT188340.1 (SKU: 102917), MT118835.1 (SKU: 102918). The threshold for detection of SARS-CoV-2 was determined by comparing the genome coverage (%) resulting from confirmed negative samples (by RT-PCR), and no-template controls and that resulting from known positive samples (by RT-PCR).

### Design of multiplex PCR panel for SARS-CoV-2 genome

A multiplex amplicon-based next generation sequencing (NGS) platform was developed to sequence the entire SARS-CoV-2 genome. Primers for 327 amplicons were designed to span the entire SARS-CoV-2 genome in tiled configuration, and alternately assigned to two separate primer pools (Pool 1 and Pool 2) to allow amplification of the whole genome while minimizing formation of short overlapping amplicons. Five primer pairs designed to target five different human housekeeping genes (*TBP, MYC, LRP1, ITGB7*, and *HMBS*) are used as control in both pools. Each forward primer additionally includes on the 5’ end, a random 10 nucleotide sequence to serve as molecular barcode. Each designed primer was checked against published SARS CoV-2 genomes as of April 15 2020, and base degeneracy was incorporated when required to achieve coverage of >99% of Asian, USA, Europe and Chinese genomes published.

### Preparation of library for NGS of SARS-CoV-2

Based on the CDC RT-PCR results, extracted nucleic acid were diluted 10-fold for samples with cycle threshold (Ct) ≤20 or undiluted for samples with Ct>20, followed by reverse transcription using High-Capacity cDNA Reverse Transcription Kit (Applied Biosystems, USA) to generate complementary DNA (cDNA). A synthetic spike-in RNA control was added to the sample just before cDNA synthesis. The RNA control comprises a nucleotide sequence which is not found in a genome or a transcriptome of the virus, and is further not found in a genome or a transcriptome of a human, and is further not found in a genome or a transcriptome of a microorganism. Each cDNA was split into two reactions for target capture and enrichment of Pool 1 and Pool 2 using Platinum SuperFi II DNA Polymerase (Invitrogen, USA) under the following thermocycling conditions: Denaturation at 98^0^C for 30s, followed by 5 to 15 cycles (5 cycles for samples with Ct <15, 10 cycles for samples with Ct 15-30, 15 cycles for samples with Ct >30) of 98°C for 1 min, 60°C for 1 min, and 72°C for 1 min, with final extension at 72°C for 5 min. At the end of the reaction, excess primers were removed by purification two times with 1.5x AMPure XP beads (Beckman Coulter,USA). Purified products were subjected to a final PCR to amplify targets and to complete the library with indexed sequencing adaptors for sequencing on the Illumina platform. Briefly, purified product was amplified with indexed P5 adapter sequence and indexed P7 adapter sequence using KAPA HiFi HotStart ReadyMix under the following thermocycling conditions: Denaturation at 98°C for 45 s, followed by 18 cycles of 98°C for 15 s, 60°C for 30 s, and 72°C for 30 s, with a final extension at 72°C for 1 min. The amplified library was purified twice with 0.7x AMPure XP beads to remove non-specific products. The quality and quantity of the sequencing library is assessed using the 4200 Tapestation system (Agilent Technologies, USA) and KAPA Library Quantification Kit for Illumina® Platforms (Kapa Biosystems Inc., USA) respectively. Paired-end sequencing (2×151bp) of the final dual-indexed libraries is performed on the Illumina platform as per manufacturer’s instructions.

### Primary sequencing data analysis

FASTQ files were processed using a custom pipeline. First, expected amplicons were identified and labeled in the FASTQ files based on the expected primer sequences in Read 1 and paired Read 2. Primer sequences and upstream molecular barcode sequences were trimmed using cutadapt, primer trimmed sequences were mapped to the reference genome SARS-CoV-2 reference genome (NCBI Reference Sequence: NC_045512.2) using bwa-mem. Molecular tag (or barcode) sequences were included in the trimmed “primer” sequences of read 1, and can be extracted given the unique structure of primer sequences in read 1. Barcode sequences were clustered based on sequence and amplicon identity, and consensus calling was done for each molecular tag (or barcode) cluster, by first performing global alignment among all associated reads using MAFFT. The consensus base in each aligned position was called by determining the majority representative base type, the percentage of which should be no less than an automatically determined threshold. The threshold is a function of the total number of reads for that barcode sequence. If no representative base could be called, the position was assigned N (as opposed to one of A, C, T, G). An overall quality score, of either 90^th^ percentile of all the quality values from the representative base type in that position (if a consensus base is found), or 10^th^ percentile of all quality values in that position (if no consensus bases is found) is assigned. The consensus reads are then written to a new FASTQ file.

### Variant calling, subgenomic RNA analysis and phylogenetic analysis

First, consensus FASTQ files from two pool 1 and pool 2 were merged to create a single FASTQ file. Consensus FASTQ reads are mapped to the SARS-CoV-2 reference genome (NCBI Reference Sequence: NC_045512.2) using bwa-mem^28^. Samtools^29^ was used to calculate depth per base and coverage metrics. Variant calling was performed on consensus BAM files using snippy and freebayes. For variants to be called, a minimum of 10x coverage is required. With molecular barcoding, the sequencing is error-free and can enable detection of quasispecies and increase confidence of variant calls due to the high quality of sequencing data.

Detection of subgenomic RNA (sgRNA) was done by analysis of split reads supported by multiple consensus reads. Chimeric reads in which non-contiguous regions of the genome are captured within a read are identified as split reads and have 1) unexpected combination of primers which resulted in the amplicon, 2) presence of junction sites at which fusion occurs and 3) presence of a junction core sequence for coronaviruses which is exemplified by the sequence – CTAAACGAAC – within the read^30,31^. The detection of canonical sgRNA was defined by the presence of the 5’ end leader sequence of the genome and downstream ORFs on the 3’end of the genome. Due to the design of the primers and full coverage inherent in the primer panel design, all canonical sgRNA can be detected. The frequency of each sgRNA was calculated as the number of split reads supporting sgRNA/number all reads supporting the location spanning the junction 3’ to the junction. For comparison across samples and across different species of sgRNA, the sgRNA split read count was normalized to the mean depth of coverage for the sample. For comparing total sgRNA, the sum of all sgRNA read counts was normalized to the mean depth of coverage for the sample.

FASTA file of the assembled genome are used to construct phylogenetic trees. The phylogenetic tree building process and parameters follows the NextStrain team’s repository at https://github.com/nextstrain/ncov^32^.

### Insert length analysis and calculation of viral fragmentation score

For each positive sample with sequencing data, SAMtools was used to capture insert sizes from alignment files with the following specifications: samtools view -f65 -F2048 SampleXYZ_consensus.bam | cut -f 1,4,9 > SampleXYZ_f65F2048.txt where f65 = filter in read paired, first in pair (Reads which are paired and insert sizes for first read in read pair and second in read in read pair will have same same insert size with “negative” length); F2048 = filter out reads with supplementary alignment (removes most subgenomic RNA reads which have part of reads with supplementary alignment and removes chimeric reads. Finally inserts of size 70 to 1000 are retained to get representation of expected insert sizes as no insert is expected to be shorter than 72 bp (design of inserts) and remove extremely long inserts.

Binning of inserts was done based on the expected and observed insert sizes into 0-150 bp (short) and 151-350 bp (long) ranges. Short and long insert counts were determined for each sample, representing short and long viral fragments, respectively. The viral fragmentation score is then derived as (number of fragments of size 151-350bp)/(number of fragments of size 0-150bp).

### Multiplex RT-PCR assay for simultaneous detection of fragments of different lengths

A multiplex three-target RT-PCR assay was designed for the simultaneous detection of two short targets of 70 bp each in the N gene region (in two separate detection channels), and one long amplicon of 240 bp in the S gene of the SARS-CoV-2 genome. An internal housekeeping control target was included as fourth target in the multiplex reaction. Briefly, extracted RNA (5 μl) was mixed with the primers/probes for the multiplexed targets with Luna® Universal Probe One-Step RT-qPCR Kit (New England Biolabs, USA) and RT-PCR was performed using recommended protocol for the Luna master mix on a BioRad CFX96. Cycle threshold (Ct) values for each target were collected from respective channels and compared.

### Statistical Methods

All datapoints are described individually with median and interquartile ranges, where appropriate. For comparison between groups, the Mann-Whitney unpaired t-test was used. Regression analysis and curve fitting was done using Prism 8.0.1.

## Results

### Detection of SARS-CoV-2 in clinical samples by whole-genome sequencing

Whole-genome sequencing was performed on RNA extracted from 155 samples corresponding to 48 individuals in this study. The samples included 37 nasopharyngeal swabs, 60 self-nasal swabs, and 58 saliva samples. An amplicon-based next-generation sequencing (NGS) panel covering the whole SARS-CoV-2 genome except the first 25 bases and 30 bases upstream of the final polyA tail was used. All samples had been previously characterized by the CDC RT-PCR assay, of which 11 (7.09%) of 155 samples were negative for SARS-CoV-2 and were included in the study to demonstrate the specificity of detection of SARS-CoV-2 by sequencing. The negative samples were from 6 cases which had at least one other matched positive sample from the same or different timepoint. Due to the high rate of positivity (92.9%) among the samples in this study, a set of 25 unrelated samples from the community confirmed to be negative by the same RT-PCR assay were also subjected to sequencing to establish a true threshold for calling positive based on the percentage of genome coverage obtained from sequencing. Additionally, 24 no-template controls (NTC) were included to demonstrate the specific detection of SARS-CoV-2 in samples, unaffected by background rates of formation of non-specific products from highly multiplexed amplicon-based sequencing. The cut-off of 1.8% genome coverage correctly classified 100% of positive cases, while 2 samples called negative by RT-PCR were just above the cut-off at 2% and 3% genome coverage, respectively (Fig. 1A.) No false positives were seen among the community negative and NTC samples. The genome coverage percentage for positive samples was closely related to the sample’s cycle threshold (Ct) value from RT-PCR assay (Fig. 1B). A large proportion of samples (62 of 144, 43.1%) had >99% genome coverage which corresponded to a median Ct value of 23.61 (inter-quartile range, IQR: 18.37-26.26). Average depth of coverage was slightly higher in saliva samples compared to upper respiratory tract (URT) samples (nasopharyngeal and nasal swabs) (p = 0.0281) (Fig. 1C), but dependent mainly on the viral load in the sample represented by the Ct value (Fig. 1D). Limit of detection studies with synthetic SARS-CoV-2 reference genomes for sequencing-based detection of SARS-CoV-2 was determined to be 50 copies/ml (Fig. S1) and expected detection of variants from synthetic reference genome was also validated using this method (Fig. S2).

**Figure 1.**
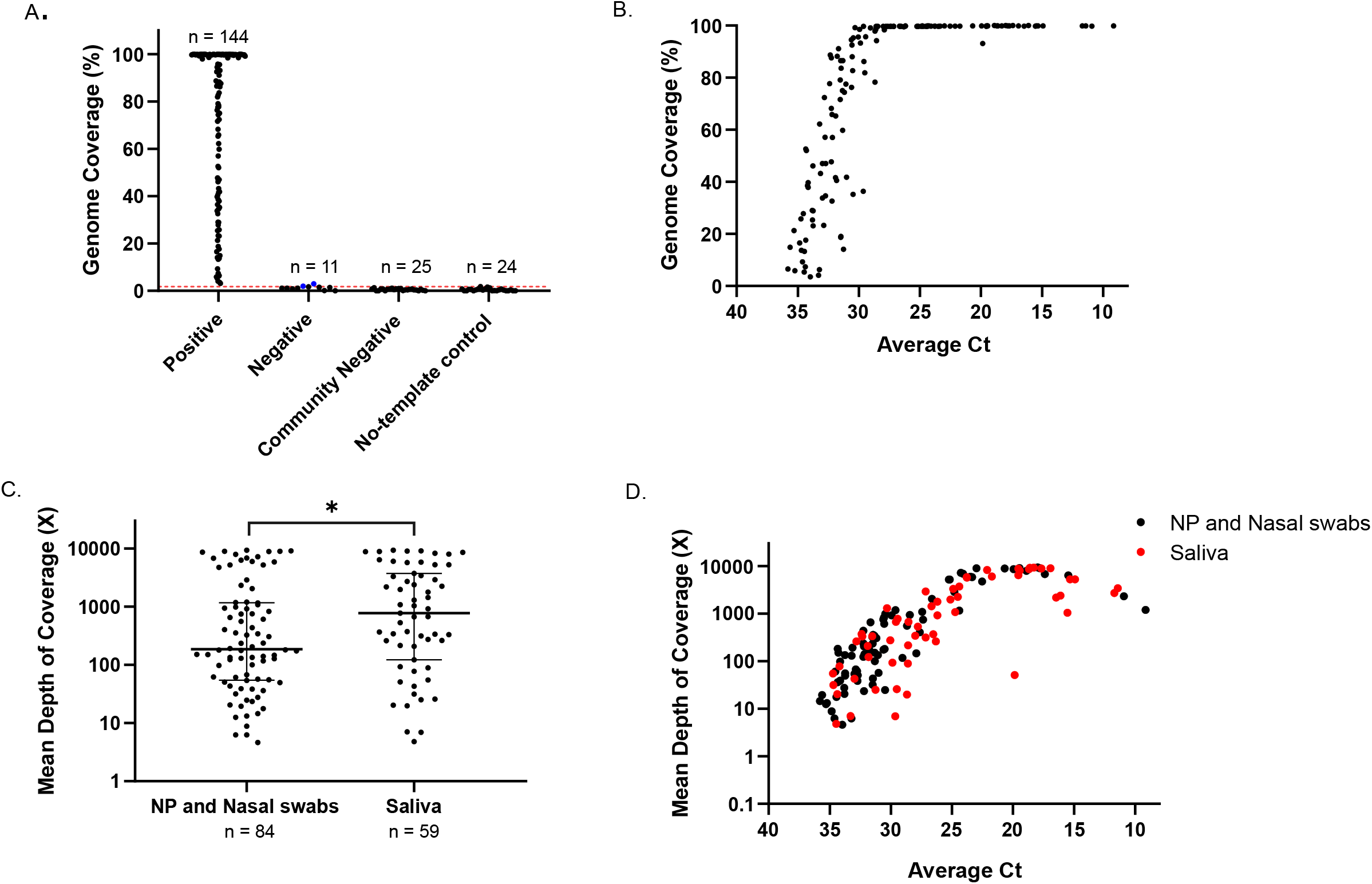
Next-generation sequencing (NGS) allows quantitative, sensitive and specific detection of SARS-CoV-2 in clinical samples. **(A)** Grouped plot showing the breadth of genome coverage for samples including positive and matched negative samples from this study, confirmed negative samples from the community, which are unlikely to have any SARS-CoV-2 and no-template control (NTC) samples run parallelly with the samples over multiple batches of library preparation for sequencing. The cut-off genome coverage for positive samples was determined to be ∼1.8% based on confirmed negative samples from the community (indicated by the red dotted line). Two of 11 samples negative by RT-PCR were just above the cut-off qualifying as positive for SARS-CoV-2 by sequencing. It remains possible that samples testing negative RT-PCR from the present study have very low-level positivity, which is at the limit of detection of the PCR assay. **(B)** The genome coverage (%) of SARS-CoV-2 by NGS (≥1x depth of coverage) is related to the original viral load in the clinical sample as measured by Ct value in RT-PCR. For clinical samples (n = 144) as Ct decreases, the % genome coverage reduces. **(C)** Mean depth of coverage is marginally higher in saliva samples (median: 186.7x, IQR: 54.33-1175x) relative to upper respiratory tract samples (URT) -nasopharygeal (NP) and nasal swabs (median: 776.8x, IQR: 122.5-3731x). *Mann-Whitney test, p = 0.0281. **(D)** Mean depth of coverage achieved is related to the viral load of the sample as measured by Ct value and decreases with increasing Ct value in both saliva and NP and nasal swabs.

### Variant analysis and phylogenetic classification

Comparative variant analysis on the SARS-CoV-2 genomes assembled from sequencing data was done for samples with >95% genome coverage (69 of 144, 47.9%) using the Wuhan/Hu-1/2019 genome as reference. Comparing samples originating from a single individual, samples from multiple timepoints, and those from differing sampling sites (upper respiratory tract or saliva), variants confidently detected (supported by >20x depth of coverage) were completely identical (Fig. 2A). This is contrast to a study in which throat swab and sputum from a patient sampled on the same day showed the presence of a unique variant in the throat swab, suggestive of independent replication in the two tissue sites^8^. Among the variants identified, a stop codon in ORF8 gene c.175G>T p.Glu59* - 28068G>T was observed in 6 samples coming from one individual, and 2 samples and one sample from two other individuals, respectively. Premature stop mutations in ORF8 have been reported among SARS-CoV-2 sequences in GenBank and GISAID initiative^33^.

**Figure 2.**
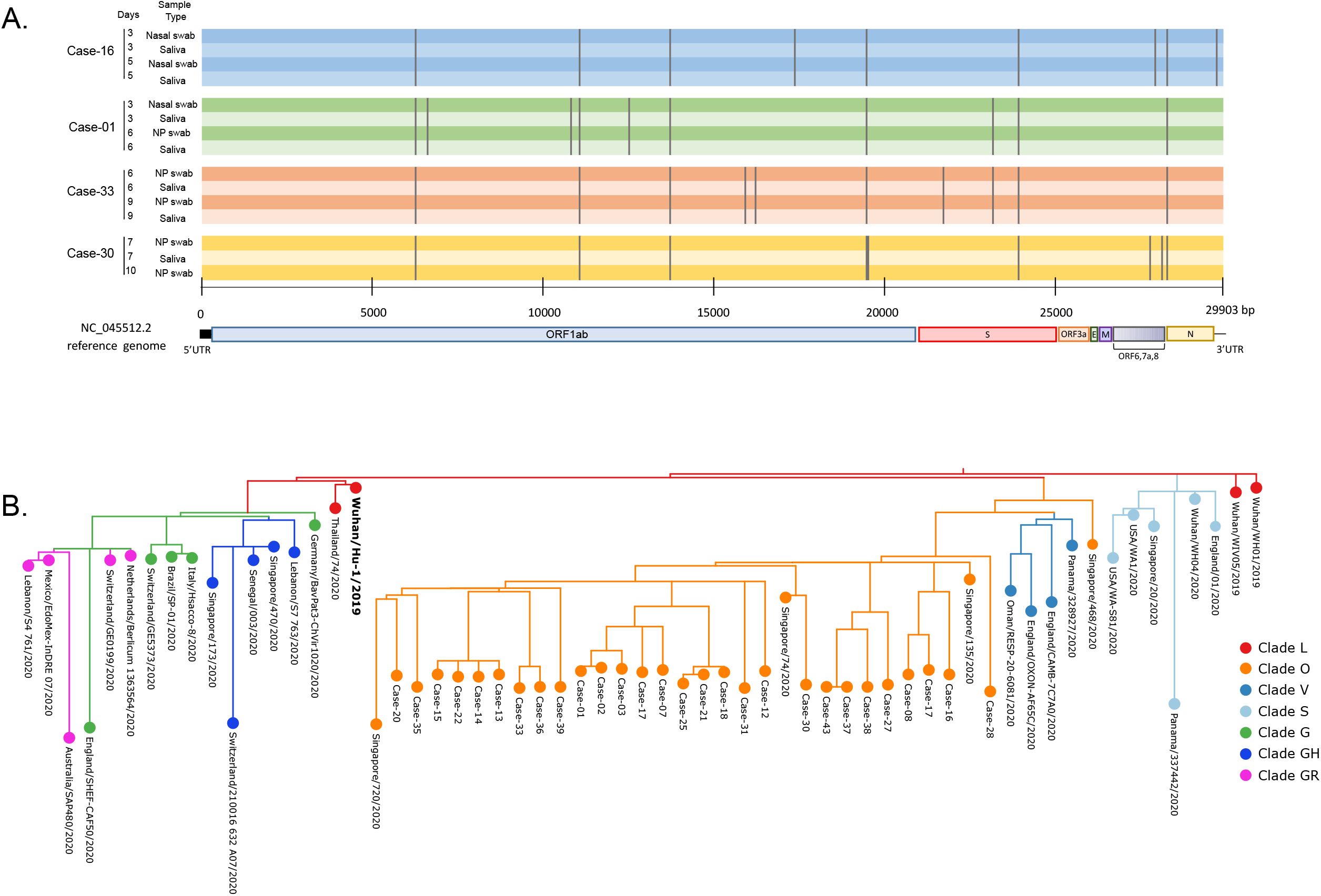
Genomic sequence analysis of SARS-CoV-2 in samples from multiple sites and over serial timepoints. **(A)** Genetic variant profiles of sequenced SARS-CoV-2 genome relative to the Wuhan/Hu-1/2019 reference from 4 representative cases with samples from multiple sites (NP or nasal swab or saliva) and over different time points since symptom onset (days). Genomic regions are depicted by shaded areas colored according to the case, and the grey lines represent the position of the detected variants. For each case with multiple samples each with >95% genome coverage (16 cases, not all depicted), identities of confidently called variants were conserved among the case’s samples. **(B)** Phylogenetic tree of SARS-CoV-2 genomes from 28 cases relative to each other and to selected samples representing 7 GISAID clades (branches colored by clades). Cases from this study all belong to GISAID Clade O (orange branch) and cluster with each other and other Singapore samples. Phylogenetic reconstruction was performed using Nextstrain (https://nextstrain.org/) and GISAID (Global Initiative; https://www.gisaid.org).

As the identity of variants among multiple samples from the same case were conserved, a single representative sequence from each case was used for the phylogenetic tree construction. A total of 28 cases (58.3%) were analyzed, as they had at least one sample satisfying the >92% genome coverage lacking stretches of N’s required for phylogenetic analysis^34^. All sequences were identical or highly similar to sequences reported in previous analyses of SARS-CoV-2 cases in Singapore, and belonged to Clade O by GISAID nomenclature^35^, and lineage B.6 by PANGOLIN system of nomenclature^36^ and with respect to each other the 28 cases fit into 3 main clusters (Fig. 2B). One Case-28 which formed a single-case branch, harboured three unique variants M c.120C>T p.Ala40Ala - 26642C>T and ORF3a c.377G>A p.Arg126Lys - 25769G>A and orf1a c.6267G>T p.Glu2089Asp - 6532G>T in four different samples collected for this case.

### Frequent detection of subgenomic RNA in clinical samples

The panel of primers for whole-genome sequencing of SARS-CoV-2 was specifically designed to also capture subgenomic RNA (sgRNA) sequences involving the fusion of the 5’ leader sequence (first 72 nt of the genome) of the SARS-CoV-2 genome with the 5’ end of each mRNA coding sequence, called the “body”. The tiled design of primer pairs and inclusion of the primer targeting the 5’ leader in both primer pools allows the capture of sequences corresponding to canonical sgRNA containing a consensus core sequence^24^. The relative abundance of each sgRNA was normalized by the mean genome depth of coverage of a sample to make comparisons across samples and sgRNA species. Among individuals with symptoms, sgRNA was found be relatively more abundant when found in URT (NP and nasopharyngeal) samples relative to saliva samples (unmatched) and was mostly detectable between -1 to 7 days since symptom onset (Fig. 3A). In both URT samples and saliva, the three most abundant sgRNA species were N, M genes and ORF3a. ORF10 was detected in only one of 68 evaluable positive URT samples, and was not detected in any of 59 positive saliva samples from symptomatic cases. This is in accordance with previous findings which found almost no subgenomic reads corresponding to ORF10^24,25,31^. Presence of sgRNA is considered an indicator of active viral replication^8,11^ with more frequent detection in samples collected <8 days after symptom onset^11^ and has been reported to persist up to 22 days^37^ after onset of clinical symptoms Among samples in the present study, the persistence of sgRNA detection was observed in samples from 8-12 days since symptom onset and up to day 13 in saliva (Fig. 3A). Samples from asymptomatic cases also had prevalent sgRNA detection in URT samples 0-3 days since diagnosis (Fig. 3B). In contrast to URT samples, in saliva from asymptomatic cases, sgRNA could be detected in up to 5-6 days since diagnosis. No sgRNA was detected at 8 days since diagnosis. Individually assessed, detection rate of sgRNA progressively decreased over time (Fig. S3). Across all types of samples and cases, when detected, the relative total amount of sgRNA (sum of individual sgRNA) did not appear to be related to the viral load of samples as measured by the Ct values (Fig. 3C). However, for samples with Ct ≥28.67 (n = 80), 61 (76.25%) had no sgRNA expression while 100% of samples with Ct <28.67 (n = 64), had some sgRNA expression, for an overall sgRNA detection rate of 57.6% in all samples (Fig. 3C). These results suggest that sgRNA is frequently detected in multiple samples types containing a minimal viral load related to Ct value in both symptomatic and asymptomatic cases. However, there is no relation between the abundance of sgRNA and Ct; following an approximate dichotomy of detection around Ct 28.7.

**Figure 3.**
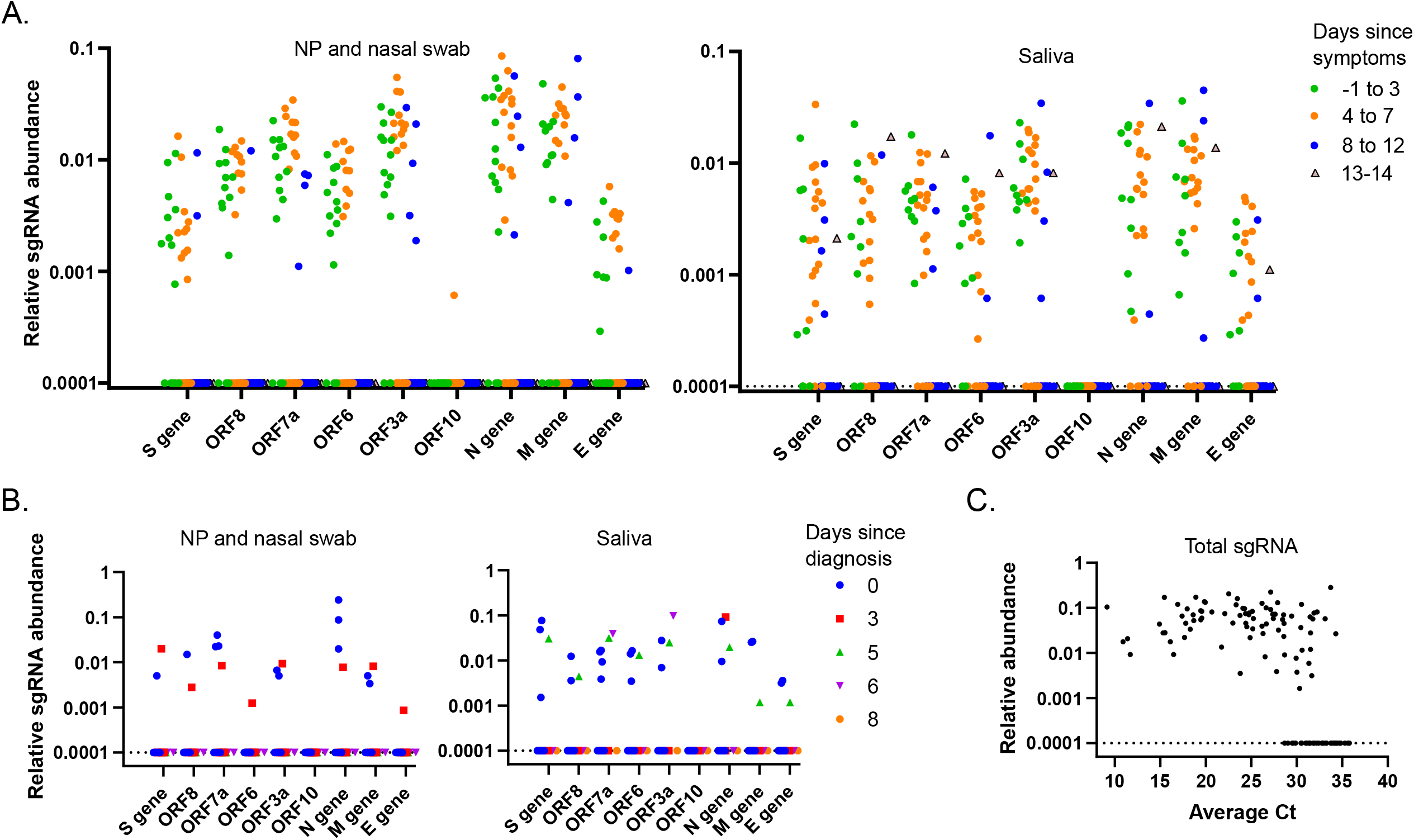
Detection of subgenomic RNA (sgRNA) in samples from symptomatic and asymptomatic cases over time. **(A)** In symptomatic cases, sgRNA species were abundantly detectable in both NP and nasal swab (URT) and saliva samples, and showed persistence up to 13 days since symptoms. Positive detection of sgRNA was largely within -1 to 7 days since symptom onset. N, M genes and ORF3a were the most abundant sgRNA in both sample types, while ORF10 was detected in only one sample. **(B)** In asymptomatic cases, sgRNA was most detectable in samples collected 0-3 days since diagnosis in both URT and saliva and was seen to persist up to 5-6 days since diagnosis in saliva. (C) Amount of total sgRNA, when detected, was not correlated with Ct value, but showed a Ct-related drop-off in detection, where 76.25% (61 of 80) samples with Ct≥28.67 did not have any sgRNA. Graph combines URT and saliva samples from symptomatic and asymptomatic cases. Zero values have been converted to 0.0001 to allow plotting on logarithmic scale and cluster on y-axes line at 0.0001.

### SARS-CoV-2 RNA fragments of differing lengths are captured by NGS

We observed that one of the consequences of tiling multiple primer pairs (in subsequence) to capture the entire known target template of SARS-CoV-2 in a highly multiplexed PCR-based NGS method, was the capture of longer inserts in addition to the inserts expected to be formed from amplicons of length 130-178 bp for which primer pairs were originally designed (Fig. S4A). The origin of the longer inserts was the formation of amplicons between a given forward primer and a reverse primer that is 1-, 2-, 3-, 4- and so on-displaced in the subsequence of primer pairs in the pool of primers (Fig. S4B). We reasoned that the more degraded the viral RNA template, the more the capture of short inserts (representing short fragments) would be favoured, due to the unavailability of intact RNA template for longer amplicons to form (Fig. S4C). Analysis of the sequencing data for the distribution of insert lengths per sample, showed that 2 to 3 distinct ranges of insert lengths could be captured per sample, and the number of sequencing reads supporting inserts in each insert length group could vary substantially between samples (Fig. 4A). Based on the dominance of inserts of lengths either 70-150 bp or 220-350 bp in sequencing data (and not other longer insert length groups), we chose these two insert length groups for further study and refer to them as “short” and “long” fragments, respectively. The cumulative number of short and long fragments (all reads mapping to SARS-CoV-2 from all potential primer combinations across the targeted genome) for samples were determined. It is noted that the sizes of the amplicons for N1 and N2 targets from the CDC RT-PCR assay initially used to characterize the samples are 72 bp and 67 bp, allowing sensitive detection of even very small fragments of virus RNA. It follows that such an RT-PCR assay would not be able to distinguish between long and short fragments that may exist in a given sample, but would give a cumulative read-out comprising all fragments. In 144 samples positive for SARS-CoV-2 by the CDC RT-PCR assay, the sequencing read counts for long and short fragments were each correlated with Ct value as expected, diminishing as the Ct value increased or presumed viral load decreased. However, the longer fragments had a steeper change in abundance relative to Ct value, become disproportionately fewer relative to short fragments in samples with low viral load reflected by later Ct values (Fig. 4B). We hypothesize, given a mass of viral RNA (a mix of intact and degraded RNA), measurable as a Ct value by an assay like the CDC RT-PCR assay with very short amplicons, that at later Ct values, previously long fragments have been converted to short fragments due to degradation of viral RNA, and contribute in turn to the observed read counts for short fragments which continue to maintain a relatively higher count from this contribution. Consequently, degradation of viral RNA more rapidly leads to the diminishing read counts for long fragments observed at late Ct values. This suggests that counting long fragments or the relative distribution of long to short fragments would provide a more accurate read-out for presence, if any, of intact viral RNA in samples.

**Figure 4.**
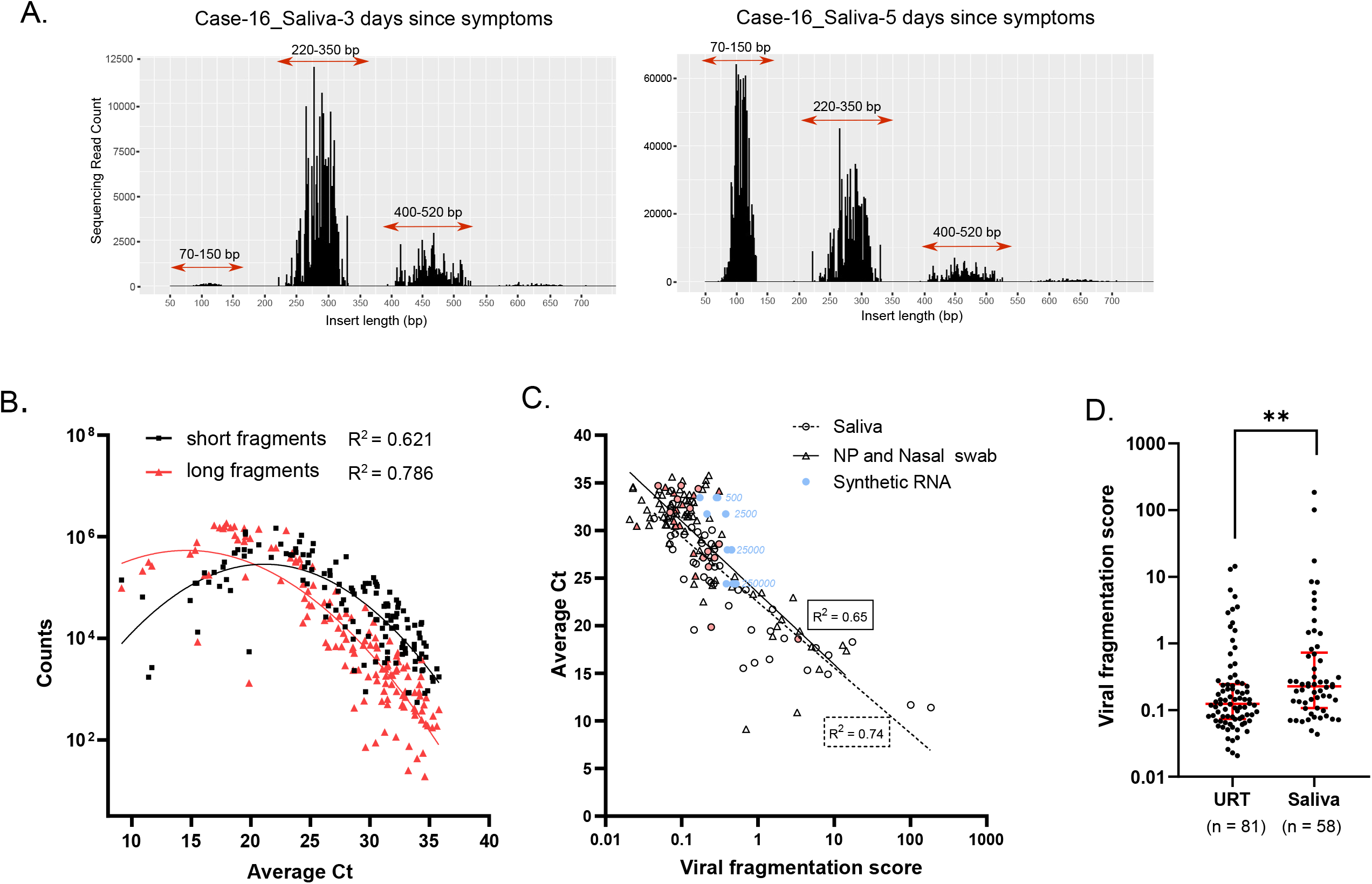
SARS-CoV-2 fragments of differing lengths can be detected by NGS as short or long inserts with varying relative abundances. **(A)** Histogram of sequencing read counts supporting insert lengths of ranges 70-150 bp, 220-350 bp, and 400-520 bp. Insert length distributions in longitudinal saliva samples from a single case are shown as examples. The relative abundance of insert lengths changes from a dominant 220-350 bp (long inserts) distribution in a sample taken 3 days from symptom onset, to a more abundant 70-150 bp (short inserts) distribution prominent in a sample taken 5 days from symptom onset. Insert lengths are representative of viral RNA fragments lengths. **(B)** Long and short fragment counts are each correlated with the Ct value of the sample measured by the CDC RT-PCR assay. Long fragments have a steeper decline in abundance, becoming differentially fewer compared to short fragments at Ct values greater than 25. URT (NP and nasal swab) and saliva samples are plotted together. **(C)** Long fragments count/short fragments count = viral fragmentation score (VFS) is moderately correlated with Ct value of a sample, in both URT and saliva samples. R2 values of correlation are shown. Pink-filled circles represent asymptomatic cases. Blue circles represent synthetic RNA samples of known integrity that generate a considerably constant VFS (median: 0.384, IQR: 0.289-0.449) across Ct values 24.4-33.45 over 500-fold serial dilution of synthetic RNA. Numbers next to blue circles are the concentrations in copies/ml of synthetic RNA. **(D)** Viral fragmentation scores are significantly higher across saliva samples compared to URT samples. Red lines represent median and interquartile ranges. **Mann-Whitney test p = 0.0035.

We devised a viral fragmentation score, VFS (long to short fragment count ratio) to capture this variability in distribution of fragment lengths. VFS showed a moderate correlation with Ct (R^2^ = 0.65 in NP and nasal swabs, and R^2^ = 0.74 in saliva), suggesting relative abundances of long and short fragments within a sample are not captured fully by its Ct value, despite detection of long fragments diminishing with increasing Ct values (Fig. 4C). For 5 samples sequencing libraries were repeated from RNA extracts and VFS were shown to be within reproducible range for the same sample (Table S1). To rule out a fully Ct value-dependent behaviour of VFS, we made use of intact synthetic SARS-CoV-2 RNA genomes and prepared sequencing libraries from serial dilutions of synthetic RNA spiked in to negative clinical matrix and extracted via routine procedures. Synthetic RNA mimics SARS-CoV-2 genome but is composed of 6 fragments of 5000 bp each representing the genome, which we take as a relatively intact starting material. For the amounts of synthetic RNA spiked in-250000, 25000, 2500 and 500 copies/ml - the Ct values measured by the same CDC RT-PCR assay were on average 24.4, 27.97, 31.73 and 33.45, respectively. Since the RNA in the serial dilutions are similarly intact, being derived from the same stock of synthetic RNA, the increase in Ct values are attributable only to the total copy numbers of RNA detectable by the RT-PCR assay. However, the VFS for the serial dilutions of synthetic RNA, do not show the same degree of decrease as the trend for increasing Ct values and ranged with a median: 0.384 (IQR: 0.289-0.449) across all 4 serial dilutions (Fig. 4C, Fig. S5). This experiment shows that factors besides total viral RNA amount measured by a Ct value (with short PCR amplicons) determine the VFS, which depends on the presence of long fragments, more of which is likely to be present in a sample with more intact. This is supported by the wide variation in VFS that was observed in clinical samples, which was higher in saliva relative to URT samples (p = 0.0035) and ranged from 0.0433 to 184.9 in saliva (n =58, median: 0.267, IQR: 0.113-0.68) and from 0.02073 to 14.38 in URT samples (n =81, median: 0.124, IQR: 0.074-0.246) (Fig. 4D). On the basis of VFS, on average, saliva samples in this study had less fragmented SARS-CoV-2 RNA. However, a relation with the disease time-course, symptomatic/asymptomatic status as an explanation for this observation cannot be ruled out. Importantly, what constitutes a high enough VFS score to be considered clinically relevant as infectious virus with intact RNA is not established. Overall, these results support the presence of a wide range of fragmentation profiles in clinical samples which is readily detectable by NGS.

### Correlation of viral fragmentation scores (VFS) with clinically relevant measures of infectiousness

As our NGS method was able to quantify relative abundances of short and long fragments representing less and more intact virus, respectively, next we determined if VFS was related to other clinical correlates of infectivity. This is particularly relevant as it is well-recognised that RT-PCR positivity alone, particularly from a test designed for short targets, does not translate to viable virus with infection potential^5,6^. We have already shown that VFS is moderately correlated with Ct value (Fig. 4C), which in itself is considered a proxy for infectious potential of SARS-CoV-2 in clinical samples based on the culturability of virus isolates^8,11,12,22,38,39^ although with widely differing Ct cut-offs. In multiple studies, no live virus has been culturable after eight days in non-immunocompromised patients^2,10,14^. The typical clinical profile of the symptomatic cases in our study was immunocompetent with presentation of mild to moderate respiratory symptoms. Compared directly, VFS and days since symptom onset was not strongly correlated in both URT and saliva samples over the full duration of the symptom onset (days -1 to day 14) (Fig. 5A and 5B). However, transposing a VFS derived from a sample with known integrity i.e. synthetic RNA (refer to Fig. 4C), subject to the same extraction and sequencing procedure as the clinical samples, we selected a cut-off of 0.382 to separate samples with intact RNA from those without. We noted that with this cut-off value, no URT samples (total 69) collected >8 days since symptoms (n= 36) qualified as having intact RNA (Fig. 5A, top left quadrant). In saliva samples (total 46), only 1 (5.26%) of 19 samples collected >8 days since symptoms had a VFS indicative of intact viral RNA (Fig. 5B). Of 37 URT samples collected ≤8 days, 15 (40.5%), and of 29 saliva samples collected ≤8 days, 16 (55%) had VFS greater than 0.382 suggesting intact RNA profiles. These results suggest about 50% of samples collected ≤8 days have relatively fragmented RNA, in accordance with observations made in previous studies where about 29%-42% of samples with high viral load inferred from low Ct or from ≤8 days did not recover live virus^9,11,22^. Among asymptomatic cases, 25 (96%) of 26 samples had low VFS below 0.382 cut-off, from 0 to 8 days from diagnosis, suggesting that viral RNA from most asymptomatic cases is fragmented (Fig. 5C). For one case, VFS 3.36 of saliva sample collected on the day of diagnosis was matched with URT samples (NP and nasal swabs) from the same case with VFS of 0.79 and 0.14, suggesting a less fragmented SARS-CoV-2 RNA in saliva even at same timepoint. Overall, VFS differed significantly between samples collected ≤8 days from symptoms and those after 8 days for cases with information on symptom onset (Fig. 5D, left), but it was difficult to find a time measure that trended with VFS for asymptomatic cases (Fig. 5D, right). For cases with multiple longitudinal samples, VFS were tracked with respect to time from symptom onset and were generally observed to be higher early between days -1 to 6 and generally decreased with time reaching levels similar to those seen asymptomatic cases through the course of the asymptomatic infection (Fig. 5E).

**Figure 5.**
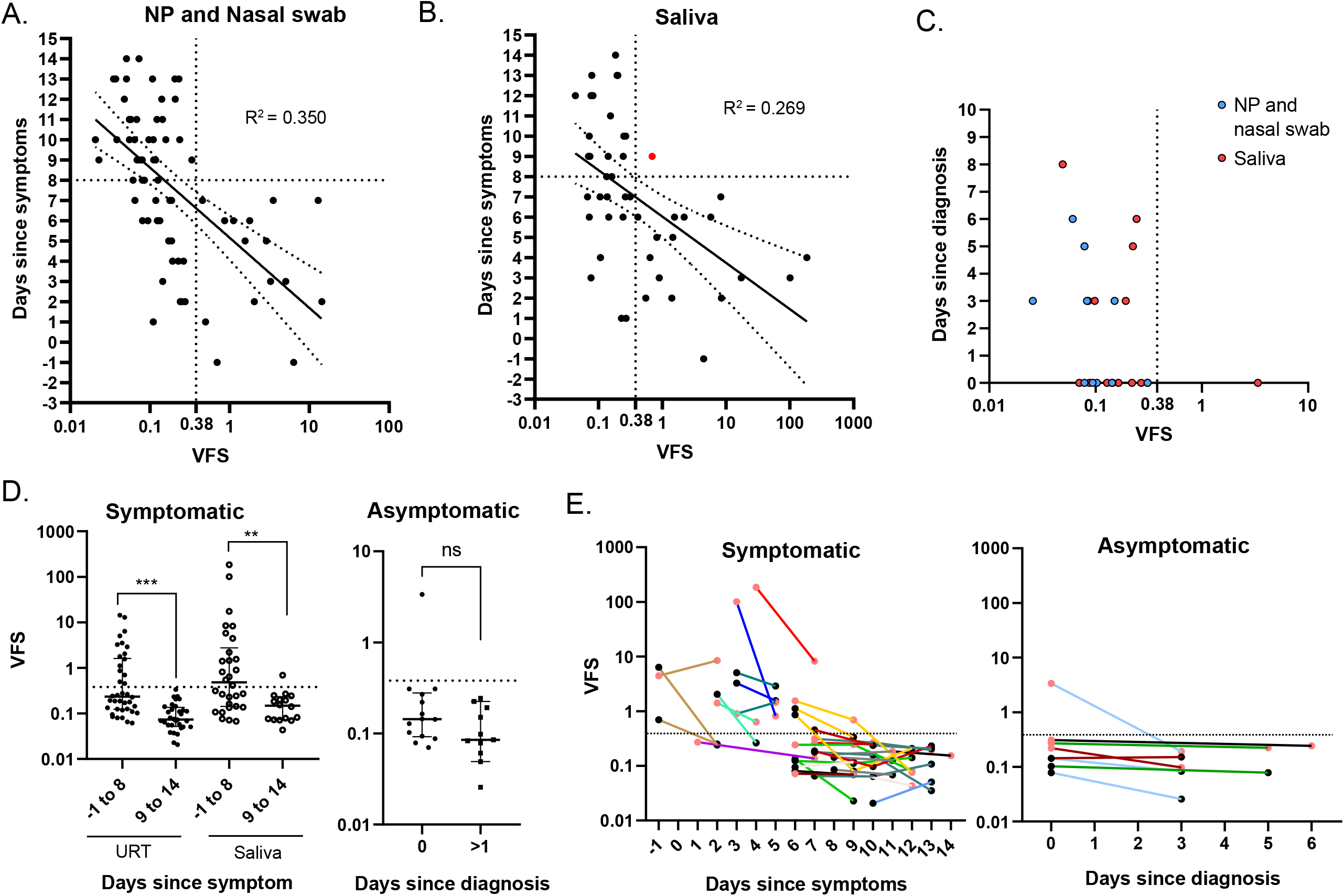
Viral fragmentation score is related to the clinical time-course of SARS-CoV-2 infection. In symptomatic cases, the relative amount of long fragments (viral fragmentation score, VFS) decreases with days since symptoms in both **(A)** URT and **(B)** saliva samples. With the exception of one saliva sample (red circle), beyond 8 days since symptoms, VFS does not exceed the cut-off of 0.38 (dotted line) estimated for intact RNA based on synthetic SARS-CoV-2 (upper left quadrants). For samples collected ≤8 days since symptoms onset, 15 (40.5%) of 37 URT samples and 16 (55%) of 29 saliva samples exceeded the cut-off (lower right quadrant). The fitted regression lines and 95% CI error are shown on graphs. **(C)** Most (96%) samples from asymptomatic cases had low VFS. **(D)** (Left) VFS differ significantly in URT and saliva between samples collected ≤8 days after symptoms onset and later samples.***Mann-Whitney test p<0.0001, **p=0.0017. (Right) No significant difference between VFS from samples collected on day 0 of diagnosis and later samples in asymptomatic cases (URT and saliva samples combined). Median and IQR lines are shown. **(E)** Matched longitudinal samples from 15 symptomatic cases (left) and 4 asymptomatic cases (right) and VFS. Each case is colored by a different line. Lines marked by pink ends are saliva samples while the rest are URT samples.

We observed no linear correlation between increasing VFS and total sgRNA abundance (Fig. 6A), but there was a significant difference in VFS of samples with any sgRNA compared to those with none, showing a dichotomous distribution (p<0.0001) (Fig. 6B). The dichotomy of distribution was related to the VFS cut-off of 0.382 determined earlier. All 33 samples with VFS ≥0.382 had some sgRNA expression, whereas 58 (54%) of 107 samples with VFS <0.382 had no sgRNA (Fig. 6A, 6B). When individual species of sgRNA were separately considered, interestingly, there was an inverse correlation with the VFS for S and N genes only, and not for other most abundant sgRNA M gene and ORF3a (Fig. 6C). No other sgRNA species showed a correlation with VFS (not shown), suggesting a specific effect of intact RNA on the abundance of S and N genes expression in clinical samples.

**Figure 6.**
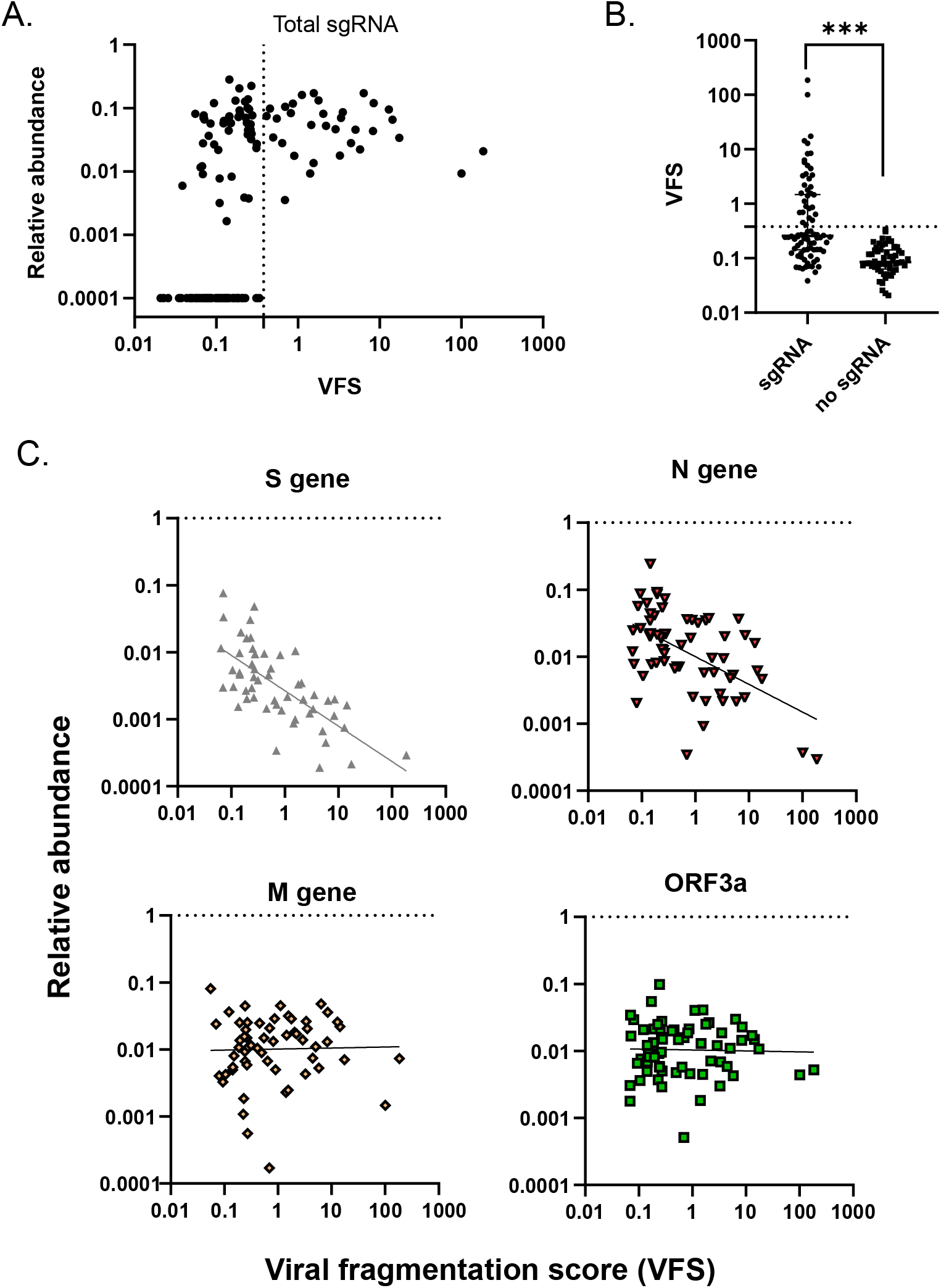
Profiles of subgenomic RNA (sgRNA) with relation to viral fragmentation score. **(A)** Across all sample types, sgRNA expression begins to tend to zero when VFS is low and a cut-off of 0.382 (dotted line) separates, 33 samples, all of which have VFS ≥0.382 and sgRNA detected, and 58 (54%) of 107 samples with VFS <0.382 having no sgRNA. **(B)** Significant difference in VFS between samples with zero or any sgRNA detected. ***Mann-Whitney test p<0.0001. Dotted line is VFS cut-off of 0.382. **(C)** S gene and N gene abundances are inversely related to the VFS, but other abundant sgRNA E gene and ORF3a are not.

Finally, we demonstrate that the viral fragmentation score (VFS) can be translated into a simple multiplexed RT-PCR assay deliberately composed of a long amplicon of 240 bp and short amplicon of 70 bp to detect the long and short fragments observed in clinical samples using NGS. In a set of 20 clinical samples, a good correlation was seen between VFS from NGS and from the difference in Ct signal of short 70 bp and Ct signal of long 240 bp amplicon, here referred to as a viral fragmentation index (VFI) (Fig. 7A). For 7 samples with low VFS, 240bp amplicon was not detectable by PCR, potentially reflecting the higher sensitivity of NGS for detection due to the interrogation of the whole genome with multiple targets, but still in line with the expectation of continued detection of short 70 bp amplicon in these samples. For 13 samples with detection of both 70 and 240 bp by RT-PCR, the delta Ct (70bp-240bp) VFI was correlated with the VFS (R^2^ =0.79, p<0.0001) (Fig. 7A, right). Over days since symptom onset, the VFI became more negative, indicating the increasingly greater abundance of short 70 bp amplicon, likely due to increasing fragmentation of viral RNA over clinical course of the disease (Fig. 7B).

**Figure 7.**
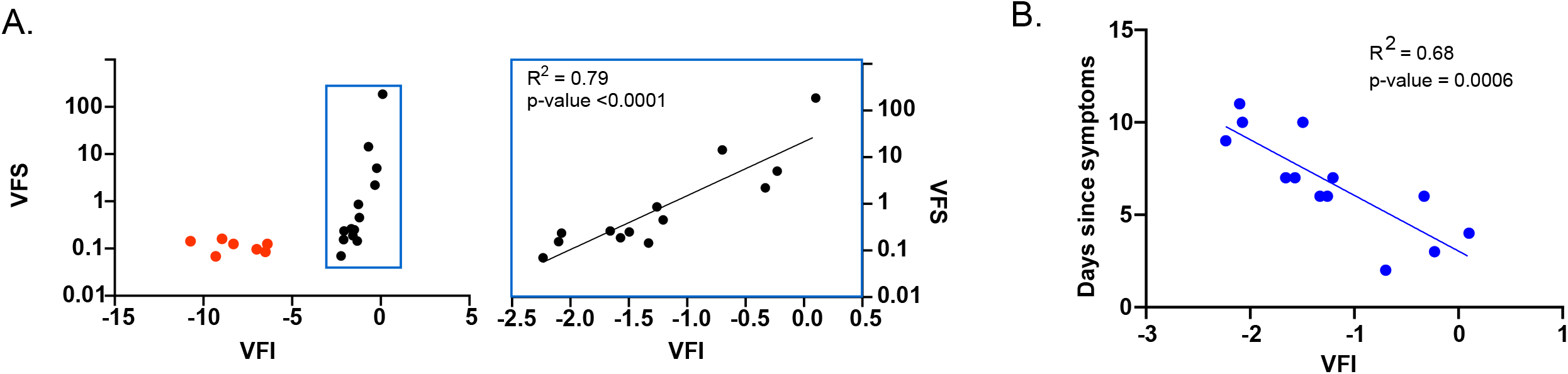
Viral fragmentation score can be captured in a multiplex RT-PCR assay for long (240 bp) and short (70 bp) amplicons. **(A)** Translation of the VFS obtained from NGS to an RT-PCR assay with calculation of difference in Ct value of short to long amplicon (Ct_70bp-Ct_240bp) for 20 clinical samples. The deltaCt value is referred to as the viral fragmentation index (VFI). (Left) Red dots are samples with no detection of 240 bp amplicon, for which Ct values are assigned to 40. (Right) Correlation of VFS and VFI for samples within blue margin is shown. **(C)** VFI from RT-PCR assay shows a correlation with days since diagnosis and for 7 samples not plotted due to non-detection of 240 bp, the days from diagnosis were ≥ 7.

## Discussion

We used next-generation sequencing (NGS) to characterize clinical samples related to the spread of SARS-CoV-2 infection in a cohort of migrant workers in Singapore, collected from different tissue sites and over serial timepoints. The design of the NGS panel allowed virus detection, genomic variant and sgRNA analysis. Further exploiting the design of the NGS panel for whole-genome sequencing of the virus, we have shown for the first time to our knowledge that NGS can be used for the detection of differential fragment lengths of viral RNA in clinical samples, potentially related to the integrity and transmissibility or infectiousness of the virus. We present a novel measure of fragmentation of viral RNA, which could be translated to a test for transmission potential of a current infection.

We show that relative to the gold-standard RT-PCR assay, NGS is a sensitive and specific method of detection of SARS-CoV-2, with 100% detection of virus in clinical samples known to be positive by RT-PCR. Additional detection of virus fragments spanning ∼3% of the virus genome in two RT-PCR negative samples, highlights that NGS may be a more sensitive diagnostic method for low levels of SARS-CoV-2. This is attributable to the targeting of the whole genome of the virus in NGS compared to RT-PCR assays targeting specific loci which may not be present among the virus fragments in a sample low levels of SARS-CoV-2. In agreement with other NGS-based detection methods^40–42^, sensitivity was high with analytical limit of detection determined to be 50 copies/ml. The detection of SARS-CoV-2 was quantitative, as the genome coverage % and the mean depth of coverage of a sample which was closely related to a Ct value from an orthogonal RT-PCR assay. As this method incorporates molecular barcodes in the primer sequences, the number of consensus reads corresponding to various regions of the genome are representative of the copy number of viral RNA fragments that are present during library preparation. Other SARS-CoV-2 NGS methods do not report similar quantitative capacity^40–42^. The quantitative feature of the method allows counting of reads to correlate with other parameters as described further on. Genomic variant analysis showed complete similarity of variants in multiple samples derived from a single case, suggesting no independent replication or generation of minor variants over time course of infection, in contrast to previous reports^8,9^. Phylogenetic analyses was possible on assembled SARS-CoV-2 genomes from 28 cases and showed that all cases were infected with virus belonging to the Clade O, lineage B.6, known to be circulating in Singapore and India.

Besides the sensitive detection of virus and identification of genomic variants, the method provided novel insights into two aspects of SARS-CoV-2 infection which have become increasingly more relevant in the context of predicting the onward transmission potential of an infection. In this work, we were able to elicit information on the presence of subgenomic RNA in clinical samples as well as the relative degree of fragmentation of the viral RNA in a sample.

Detection of sgRNA has been used as a proof of viral replication in infected cells and decline over days 10-11 from infection^8^. sgRNA detection outlasted successful virus culture and was a poor predictor of successful virus culture^14^, or showed moderate agreement with virus culture, and was detected in 18 (81.8%) of 22 specimens collected <8 days after symptom onset and in 1 (9.1%) of 11 specimens collected ≥9 days after symptom onset^11^. Due to the prolonged detection of sgRNA in clinical samples including up to 22 days since start of illness, sgRNA may not be a marker of active replication but remain detectable in clinical samples due their stability^37^.

In this study, abundant sgRNA was detected in clinical samples including prolonged detection in a saliva sample taken 13 days from illness onset from a symptomatic case. In samples with high viral loads, sgRNA was consistently detected, but became undetectable in ∼75% of samples with low viral loads (Ct≥28.67), suggesting that both viral load and sgRNA stability account for its detection. This study, to our knowledge, is the first one to demonstrate direct sgRNA detection in samples from immunocompetent asymptomatic individuals^43^ although at lower levels than symptomatic cases, in line with the non-zero but reduced transmission potential of asymptomatic carriers^44^.

It is increasingly becoming apparent that positive detection of virus RNA by RT-PCR is unrelated to presence of infectious virus, as viral RNA shedding can persist even after resolution of clinical symptoms of both mild and severe disease^8–16^. While more ill patients have generally longer detection of RNA, persistent positivity by PCR is seen in patients with mild illness as well as in asymptomatic cases^5^. A better measure of infectiousness is the culture of live virus from clinical specimens as successful virus growth is reliant on the presence of intact whole virions with complete RNA genomes. By this measure, samples taken more than 8 days since symptom onset in immunocompetent patients, do not have infectious virus as no live virus could be cultured from them^8,11,22^. In immunocompromised patients or those with severe illness, live virus could be isolated up to 14 days^12^, 20 days^14^ and much longer in severely immunocompromised patients^45^. Ability to culture live virus has been shown to be similar in asymptomatic and symptomatic indivduals ^46–48^. Some studies have correlated viral loads measured by Ct value with ability to culture live virus^8,11,14,22,38,46^, and no viral growth was reported based on a Ct value cut-off (varying between >24 and ≥35)^22,38,48^. Intriguingly, a number of studies have reported limited correlation between Ct value (or viral load) and success of virus isolation^49,50^. Samples with low Ct values (Ct<23), constituted 28.6% of samples with unsuccessful viral culture ^50^, and conversely, samples with low viral loads (Ct >35) could harbor viable virus^49,50^.

The observation that not all samples with high viral loads generate live virus, suggest that factors other than viral genome copies are important. The topmost factor would be viral integrity, whether for a small or large viral load. Only the full-length intact viral RNA would represent the virus fragment that is capable of infection. It is understood that the processing of sample in the laboratory (extraction, enzymatic reactions) would lead to some degree of fragmentation, and the use of primer-limited amplicons would artificially limit fragment lengths but relative to a sample with highly fragmented viral RNA, a sample with intact viral RNA would contain more long fragments relative to short fragments.

In this work we have shown for the first time that NGS data can not only provide total viral RNA abundance information, it can also characterize the viral RNA fragments by length. This was possible due to combination of consensus read counting (related to the original copy numbers of virus RNA in a sample undergoing library preparation) and the tiled configuration of the primer panel which could capture long fragments, if present. In other words, the more intact or full-length RNA that is present in a sample, the more long fragments would be captured by the method. The relative abundances of long and short fragments was converted to a viral fragmentation score (VFS), representing the relative integrity of the virus RNA in the sample and was shown to be related to the viral load measured as Ct value by an RT-PCR assay. It is important to highlight that the CDC RT-PCR assay targets particularly short amplicons of about 70 bp, which means it would accurately detect intact RNA when present, but would continue give an abundant signal even when largely fragmented RNA was present in a sample. At early Ct values (representing abundant viral load), the relative abundance of long fragments were as much as 100-185 times more than short fragments (very high viral fragmentation score) and could not have resulted simply as a result of the specific sequencing method.

To further address if the observed relative abundances were simply a function of the copy number of template subject to sequencing, irrespective of its starting length, we showed using synthetic SARS-CoV-2 RNA, that for relatively intact RNA template molecules the VFS remains constant even over 500-fold reduction in starting copy numbers. An empirically determined cut-off from synthetic RNA applied to clinical samples was able to separate all samples collected >8 days since illness onset as having predominantly fragmented RNA. This correlated very well with the failure to culture live virus from clinical samples collected after 8-9 days of illness in immunocompetent patients ^8,11,22,38,47,51^. Conversely, based on the cut-off, about 50% of case with VFS <0.382 (hence lesser intact RNA) were collected ≤ 8 days from symptom onset. This fraction mirrors those observed previous studied with about 29%-42% of samples high viral inferred from low Ct or from ≤8 days did not recover live virus^9,11,22^.

Another variable to consider is the sample type. Viral load and duration of positivity tends to be greater in lower respiratory tract samples (sputum) compared to throat and nasal swabs^5,52^. Accordingly, prolonged ability to culture virus has been reported for samples from severely ill patients with mostly lower respiratory tract samples^14^, compared to the success rate in upper respiratory tract samples^46^. In the present study, upper respiratory tract samples (nasopharyngeal and nasal swabs) and oropharyngeal saliva samples were compared. Oropharyngeal saliva contains secretion of the salivary glands mixed with sputum from lower respiratory tract^53^, hence is more akin to lower respiratory tract samples. In line with this, we observed higher VFS (more long fragments) in saliva samples on average compared to URT samples.

One study has looked at the potential of RT-PCR assays to determine the integrity of viral RNA and suggested that strong correlation between the levels of detection of multiple amplicons spanning the intact genome is indicative of viral integrity (and demonstrated this with live virus culture)^39^, as opposed to non-correlated levels. In the light of findings in this study and especially with the evidence that both high viral load samples and low viral load samples can produce unexpected live virus culture results, another direct measure of viral integrity is urgently needed.

Despite the large body of knowledge surrounding viral infectiousness, several gaps and hurdles remain. First, Ct value as a surrogate measure of infectivity is variably correlated with success of viral culture and multiple RT-PCR assays with Ct scales that are not directly comparable are in clinical use (evidenced from the dramatically different Ct cut-offs derived based on culture positivity ^22,38,48^). Second, typical RT-PCR assays are designed to detect short target templates and will readily amplify small amounts of fragmented viral RNA, precluding any consideration of integrity of the viral RNA. Third, duration since symptom onset has been shown to correlate with success of viral culture, however, all measures suffer from recall bias ^22,46,54^. Fourth, the viral culture test is labor intensive and requires the Biosafety Level 3, which precludes it from being established in all diagnostic laboratories, and suffers from variations in accuracy and permissiveness of cell lines. Accurate measurement of infection potential becomes particularly relevant as vaccines against SARS-CoV-2 that immunize against COVID-19 disease become available, but transmission capacity of an immunized individual remains unknown^55,56^.

We emphasize that the viral fragmentation scores (VFS) determined in this study are relative and post-laboratory processing and no determination of absolute fragment lengths contained in a sample has been attempted. The viral culture assay is the closest surrogate of this measure. Further work to correlate the insert length or an RT-PCR-based measurement of integrity to virus culture is necessary to further these findings. There are important limitations to this study. The use of the exact cut-off of VFS to categorize samples into clinical groups may not universally apply. Nonetheless, the basic premise of the argument that more intact virus would reflect in more longer fragments in a clinical sample would universally apply. As the study’s main conclusions are based on NGS, with some comparison to RT-PCR results, without evidence from virus culture studies it cannot be ruled out that some of the observed infectivity measures are correlated and result from a feature of the specific NGS method.

In conclusion, we have applied NGS to comprehensively characterize longitudinal samples collected from different sites. NGS is an enabling tool that provides sequence-related information for which it is primarily designed, and also information from size and length dimensions. Based on this, we identify fragment length differences among clinical samples which are correlated to clinical features of infectiousness of SARS-CoV-2, quantification of which could be incorporated as relevant and straightforward measure to determine infection potential.

## Data Availability

The datasets generated during and/or analysed during the current study are available from the corresponding author on reasonable request.

## Acknowledgements

We would like to thank all the medical laboratory technologists at Lucence service laboratory for their hard work and support which made this study possible.

