## Supplementary Data for "A Viral Fragmentation Signature for SARS-CoV-2 in Clinical Samples Correlating with Contagiousness"

**Figure S1. Analytical Limit of Detection of the SARS-CoV-2 by NGS.** The analytical sensitivity (or the Limit of Detection, LOD) of the method was determined by serial dilution of SARS-CoV-2 transcript of known copy number (Twist Synthetic SARS-CoV-2 RNA Control 2, (MN908947.3) – Cat # 102024). Dilutions of 2500 copies, 500 copies, 250 copies, 50 copies, 25 copies per ml were spiked into pooled negative patient clinical matrix (nasopharyngeal swab viral transport medium). Triplicate extractions were performed for each dilution and RNA was extracted using 0.2 ml of sample. Libraries were prepared for evaluation by the NGS test, and the detection rate for each spike-in copy number was calculated. Based on the established threshold of 1.% coverage for positive detection (>6 amplicons), the LOD was determined to be 50 copies/ml with a 100% detection rate in 20 replicates.

| SARS-CoV-2 (copies/ml) | Replicates | Detection Rate | Percentage positive |
|---|---|---|---|
| 250000 | 3 | 3 | 100% |
| 25000 | 6 | 6 | 100% |
| 2500 | 5 | 5 | 100% |
| 500 | 6 | 6 | 100% |
| 250 | 9 | 9 | 100% |
| **50** | **20** | **20** | **100%** |
| 25 | 12 | 9 | 75% |
| 0 | 10 | 0 | 0% |

**Figure S2. Validation of detection variants in the SARS-CoV-2 genomes using NGS.** Synthetic genomes corresponding to characterized SARS-CoV-2 genomes with accession IDs LC528232, MT007544, MT106054, MT118835 and MT188340 were converted to libraries and sequenced, and variant analysis was done using the MN908947 genome as reference. The variants identified by the method and the corresponding Allelic Depth at the position of the variant is shown. DP : total depth

| Genome | Gene | Variant | Allele Depth |
|---|---|---|---|
| LC528232 | - | NONE | |
| MT007544 | orf1b | synonymous_variant c.5598T>C p.Pro1866Pro | DP:637 C:637 T:0 |
| MT007544 | S | missense_variant c.741T>G p.Ser247Arg | DP:538 G:538 T:0 |
| MT007544 | ORF3a | missense_variant c.752G>T p.Gly251Val | DP:2180 T:2180 G:0 |
| MT007544 | | intergenic_region n.29750_29759delCGATCGAGTG | DP:405 A:405 ACGATCGAGTG:0 |
| MT106054 | orf1a | synonymous_variant c.8517C>T p.Ser2839Ser | DP:458 T:458 C:0 |
| MT106054 | orf1b | synonymous_variant c.5136T>C p.His1712His | DP:1245 C:1245 T:0 |
| MT106054 | orf1b | synonymous_variant c.5508T>A p.Val1836Val | DP:686 A:686 T:0 |
| MT106054 | orf1b | missense_variant c.5708A>C p.Asp1903Ala | DP:272 C:272 A:0 |
| MT106054 | ORF8 | missense_variant c.32C>T p.Thr11Ile | DP:6716 T:6716 C:0 |
| MT106054 | ORF8 | missense_variant c.251T>C p.Leu84Ser | DP:1584 C:1580 T:4 |
| MT106054 | N | frameshift_variant c.366delC p.Tyr123fs | DP:2978 T:2969 TC:0 |
| MT106054 | N | synonymous_variant c.822C>T p.Phe274Phe | DP:2527 T:2527 C:0 |
| MT118835 | | NONE | |
| MT188340 | orf1a | synonymous_variant c.249T>C p.His83His | DP:1027 C:1027 T:0 |
| MT188340 | orf1b | missense_variant c.3943C>T p.Arg1315Cys | DP:250 T:250 C:0 |

**Figure S3. Total sgRNA and individual sgRNA detection generally decreases with days since symptoms onset in symptomatic cases.** Detection rate of any sgRNA becomes rarer over time in **(A)** NP and nasal swab samples, and **(B)** saliva samples.

A.

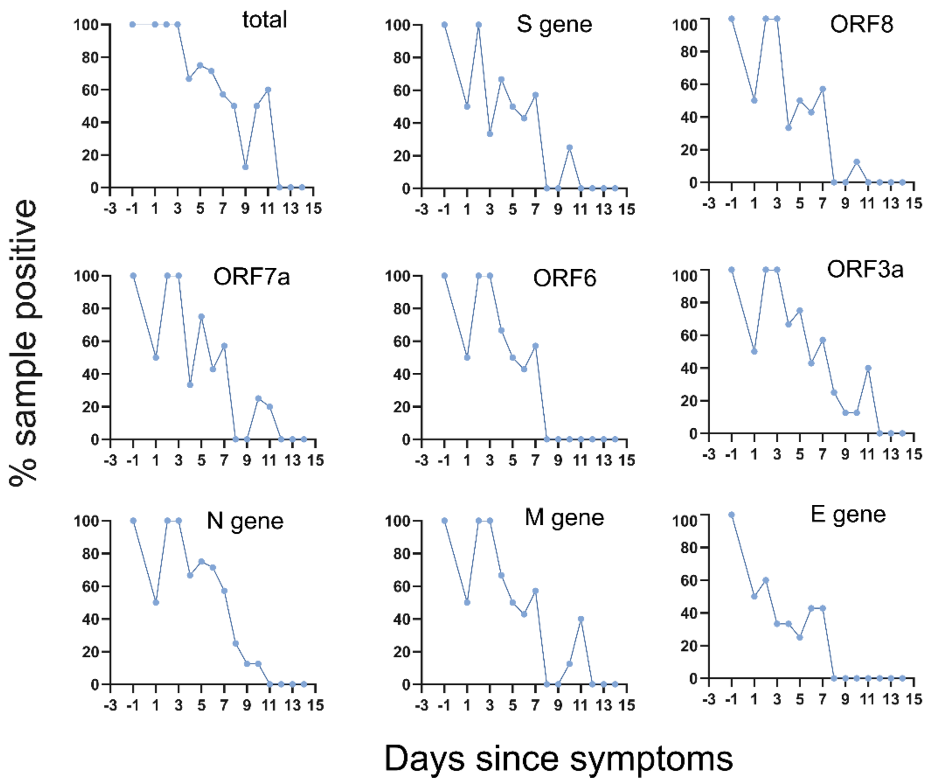


B.

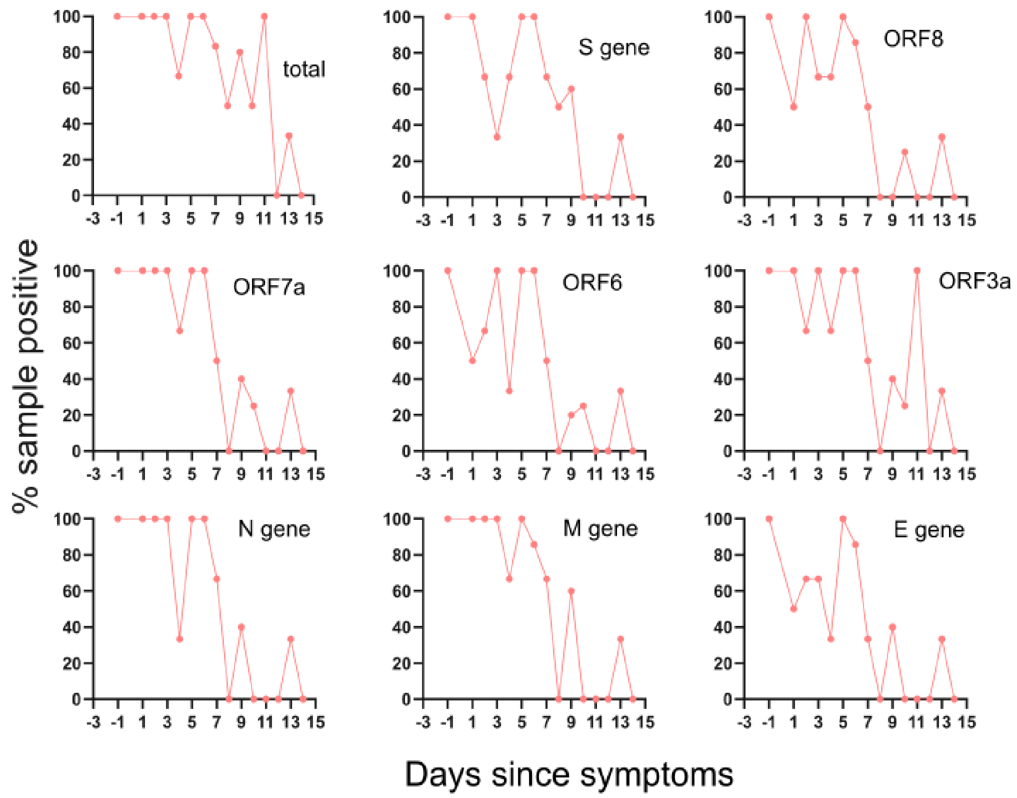

**Figure S4. Design of a highly multiplexed PCR-based NGS assay allows capture of variable lengths of RNA fragments. (A)** The definition of amplicon and insert in a PCR and multiplex PCR-based NGS assay. **(B)** Illustration of the origin of inserts of of variable lengths by amplicon-based sequencing. Primer pairs in subsequence are shown from primer 51 to 61. Inserts of length 93bp and 103bp are formed as a result of the expected interaction between 51-F and 51-R and between 57-F and 57-R. If the RNA template is intact, longer inserts can also be captured by formation of products as an example, between 51-F and 53-R and between 57-F and 59-R. These inserts would have lengths of 291 and 303 bp. Potentially, other primers in subsequence and still further from one another in productive orientations (example 51-F and 55-R) can capture still longer inserts of length 464bp. **(C)** Conversely, in samples with greater degree of fragmentation or low viral RNA amounts the shorter inserts would be favourably formed, due to the lack of long templates to be primed by distant primer pairs.

A.

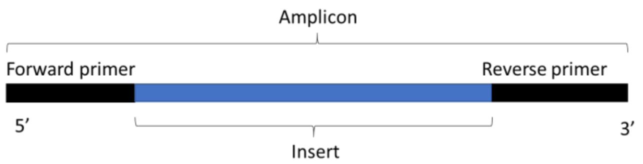


B.

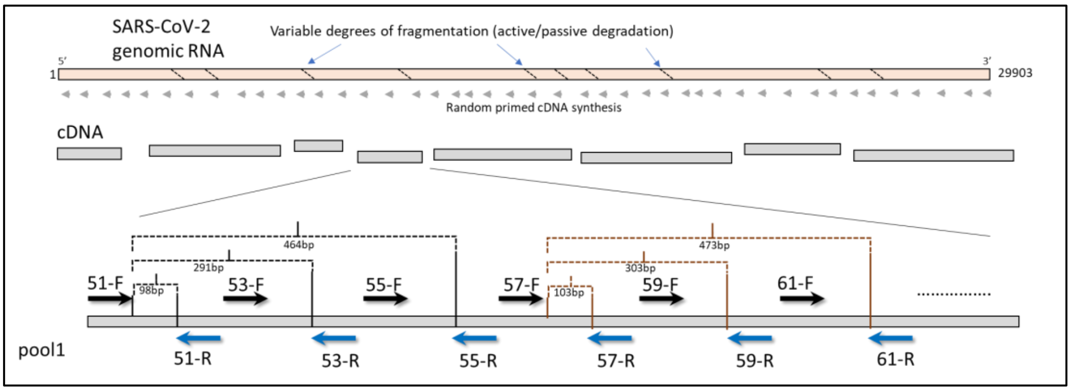


C.

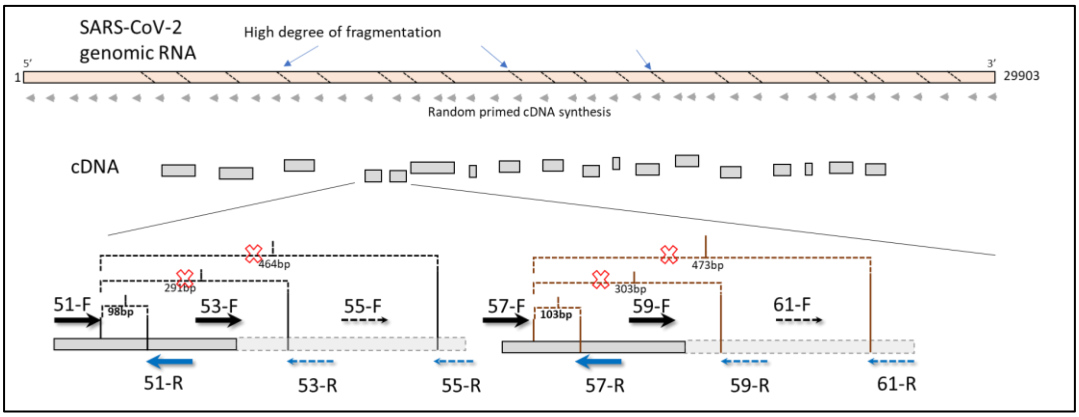

**Table S1. Reproducibility of viral fragmentation score measured by NGS in 5 clinical samples.**

| | Replicate 1 | | | Replicate 2 | | |
|---|---|---|---|---|---|---|
| Sample | Viral fragmentation score | Short fragment count | Long fragment count | Viral fragmentation score | Short fragment count | Long fragment count |
| Case-18-S2 | 0.90 | 195085 | 175475 | 1.58 | 67735 | 107331 |
| Case-36-S1 | 0.41 | 1472744 | 608118 | 0.70 | 903115 | 630520 |
| Case-39-S1 | 2.21 | 483027 | 1066185 | 5.91 | 190638 | 1125903 |
| Case-04-S1 | 14.38 | 105394 | 1515917 | 20.75 | 50176 | 1041099 |
| Case-37-S1 | 0.11 | 436845 | 46214 | 0.12 | 369195 | 45264 |

**Figure S5. Synthetic SARS-CoV-2 RNA samples in serial dilutions have expectedly increasing Ct but stable viral fragmentation score (VFS).** Synthetic RNA has a known degree of integrity of 5000 bp fragments. Sequencing libraries of dilutions of the synthetic RNA had VFS which did not decrease with increasing Ct (VFS: 0.384, IQR: 0.289-0.449). Mean values of three replicate experiments are shown. Ct values for the same serially diluted samples obtained by the CDC RT-PCR assay varied from 24.4-33.45 reflective of the range of 500-fold difference between the highest and lowest copy numbers used.

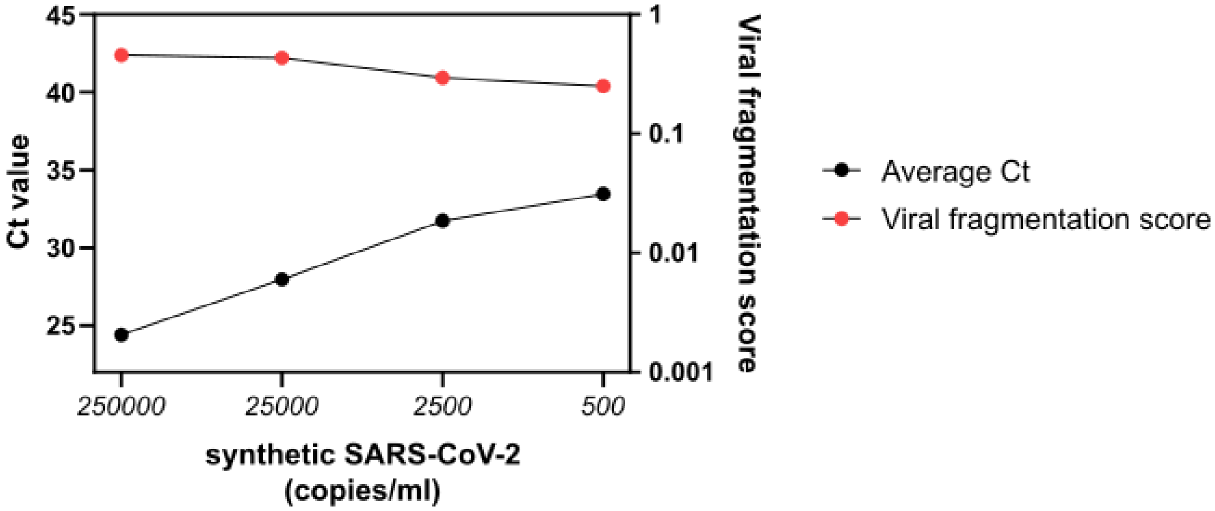
